# Underuse of hepatocellular carcinoma surveillance in patients with cirrhosis: a nested cohort study

**DOI:** 10.1101/2025.09.16.25335849

**Authors:** Pascal Caillet, Fréderic Balusson, Nathalie Ganne-Carié, Emmanuel Oger, Charlotte Costentin, Olivier Ganry

## Abstract

**Objectives:** Hepatocellular carcinoma (HCC) is the fourth leading cause of cancer-related death worldwide. Most cases occur in patients with an underlying cirrhosis. The French national guidelines recommend semiannual abdominal ultrasound surveillance for early HCC detection in patients with cirrhosis. The primary goal of our retrospective cohort study was to evaluate compliance with this recommendation.

**Methods:** We used 2007–2016 general public health insurance program (Régime Général) data from the French National Health Data System (Système National des Données de Santé, or SNDS). Included patients were 18 to 75 years old, diagnosed with liver cirrhosis between 2009 and 2013, and underwent their first ultrasound >4 months after their index date. The number of annual ultrasounds was recorded over a 3-year follow-up period. Compliance was defined as having had at least 2 ultrasounds per year over the follow-up time.

**Results:** Among the 66,464 patients included in the analysis, surveillance was optimal (no year with <2 ultrasounds) in 5,082 patients (7.6%), suboptimal (one year with <2 ultrasounds) in 3,928 (5.9%), and failed (remaining cases) in 57,454 (86.4%). Older age, male sex, a high Charlson index, frequent gastroenterologist/hepatologist visits, and viral etiology were associated with better surveillance, whereas low socioeconomic status, despite France’s universal health coverage, was linked to failed surveillance.

**Conclusions:** In French patients with cirrhosis, most of cancer surveillance is failing when considering recommendation in vigor. In order to improve surveillance, a better understanding of the social determinants of health equity is needed.

## Introduction

Hepatocellular carcinoma (HCC) is the sixth and third leading component of cancer incidence and cancer death worldwide [1,2]—behind lung, breast, colorectal, prostate, and stomach cancers—and was responsible for the death of 781,631 patients in 2018 [3]. More than 70% of cases occur in Asia, 10% in Europe, 8% in Africa, and nearly 10% in the Americas. The epidemiology of HCC has changed over the past 20 years owing to fluctuations in the prevalence of known risk factors. In 80% of cases, HCC develops within cirrhotic livers [4]. Patients with cirrhosis from any etiology typically have a ∼2% annual risk of developing HCC [5]. Once major risks in industrialized countries, viral hepatitis B and C have been declining since the introduction of highly effective antiviral treatments and the widespread distribution of the hepatitis B vaccine. In contrast, the burden of alcohol-related cirrhosis has increased, as has the incidence of cirrhosis related to nonalcoholic steatohepatitis (NASH), which mostly occurs in overweight or diabetic patients [2]. Thus, while the leading etiology of HCC worldwide is viral, alcohol-related cirrhosis and nonalcoholic fatty liver disease are the most common causes in Western countries such as France [6].

Despite advances in therapeutic management, overall mortality associated with HCC remains very high in France. HCC is among the seven cancers with the worst prognoses: estimated median survival is ∼8 months [2]. In France, the 1- and 5-year standardized net survival rates for liver cancers diagnosed in 2015 (90% of which were HCCs) were 48% and 18% respectively [7]. Survival is highly correlated with the cancer stage at diagnosis. Survival in HCC is tumor-stage dependent, with 5-year survival expected only in patients who are diagnosed at an early stage and undergo potentially curative treatments [8]. However, studies suggest that as few as only <20% of patients are diagnosed at an early stage eligible for curative options [9]. Semi-annual ultrasound screening is associated with early diagnosis, curative treatment, and survival in patients with cirrhosis [10]. Thus, standardized and effective surveillance programs for patients with cirrhosis are urgently required.

European guidelines have consistently over the past decade recommended semiannual HCC surveillance via abdominal ultrasound for all patients at high risk of HCC, including those with cirrhosis [11]. However, compliance with surveillance guidelines is low and there are significant disparities between countries and cirrhosis etiologies. Reported percentage of patients receiving monitoring per current recommendation ranges from 14% [12] to 52% [13]. Compliance in the US is usually lower than that in Asian countries [14]. It has been shown that compliance with surveillance guidelines is greater in patients with liver disease linked to hepatitis B or C [15], when follow-up is managed by a specialist [16], or when patients have significant comorbidities [17].

In France, data on HCC surveillance are scarce [6] and come from studies with small populations [17] or focused on patients with cirrhosis related to hepatitis B or C [10]. A detailed large-scale review of HCC surveillance in France is required. The primary objective of this study was to evaluate compliance with surveillance guidelines for early HCC detection in clinical practice in patients with cirrhosis in France. The secondary objective was to identify factors predicting failed surveillance.

## Materials and Methods

We conducted a retrospective cohort study using the French National Health Data System (Système National des Données de Santé, or SNDS), considering data from 2007 to 2016 for the general public health insurance program (Régime Général). The SNDS combines several previously existing databases: the nationwide claims database of the French public health insurance administration (Système National d’Informations Interrégimes de l’Assurance Maladie, or SNIIRAM), the national hospital database (Programme de Médicalisation des Systèmes d’Information, or PMSI), and the national death registry (Centre d’Épidémiologie sur les Causes Médicales de Décès, or CEPIDC) (Fig 1).

**Figure 1.**
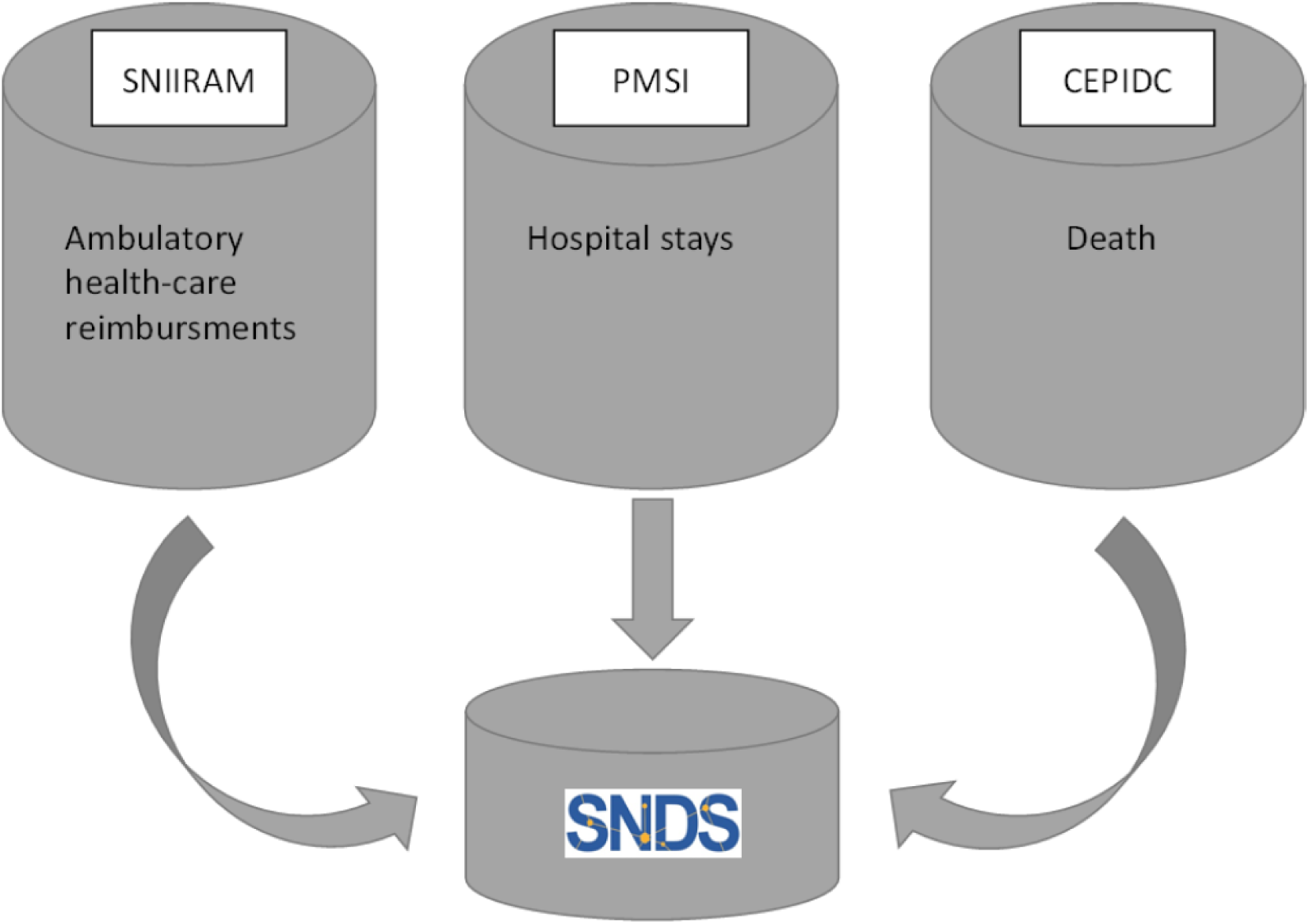
Simplified structure of SNDS. SNDS, Système National des Données de Santé (French National Health Data System); SNIIRAM, Système National d’Informations Interrégimes de l’Assurance Maladie (nationwide claims database of French public health insurance administration); PMSI, Programme de Médicalisation des Systèmes d’Information (national hospital database); CEPIDC, Centre d’Épidémiologie sur les Causes Médicales de Décès (national death registry).

The SNDS covers more than 98% (66 million people) of the French population, from birth (or immigration) to death (or emigration), irrespective of occupational changes or retirement. It is an anonymized record of individual patients’ medical encounters, hospital diagnoses, drug dispensations related to outpatient medical care claims (including all reimbursed drugs), information from hospital discharge summaries, and the dates of death. SNDS is extensively used for epidemiological research, including cirrhosis epidemiology [18–20].

The inclusion period was 2009–2013. Data from 2007 and 2008 were used to facilitate the application of the exclusion criteria by identifying incident cirrhosis. For each patient, the follow-up lasted for ≥3 years, ending no later than December 31, 2016.

The study population included patients ages 18 to 75 with an incident diagnosis of cirrhosis during the inclusion period. Diagnosis of cirrhosis was defined as the presence of at least one ICD-10 code related to cirrhosis or its complications and was associated with hospital stay during the inclusion period (S1 Table). Patients were only included if they presented with an ICD-10 code related to cirrhosis handled during hospitalization, if the date of their first cirrhosis-related ICD-10 code fell within the inclusion period, and if they were not diagnosed with cancer (including HCC) within the two years preceding inclusion.

The primary objective of this study was to assess compliance with surveillance recommendations in patients with cirrhosis. The event of interest was reimbursement for abdominal ultrasound, as indicated by the codes recorded in the SNDS (S2 Table). Patients for whom follow-up was <4 months after the inclusion date were excluded from the analysis, based on the hypothesis that these procedures may have been performed for the purpose of diagnosing cirrhosis, rather than for HCC surveillance. All scans that occurred during the first four months of follow-up were discarded.

The secondary objective was to evaluate the sociodemographic and medical factors that are believed to be associated with optimal management. Accordingly, among the additional data retrieved were patient age at the time of study inclusion, gender, universal health coverage (Couverture Maladie Universelle, or CMU) beneficiary status, indicative of low socioeconomic level; type of health facility where cirrhosis was diagnosed or managed (i.e., public university hospital, other public hospital, or private clinic); Charlson index, the number of times a year the patient was seen by gastroenterologists/hepatologists (during private appointments or through hospitalization in a GI department); and etiology of cirrhosis (i.e., alcohol-related, viral, or unknown).

This study was approved—registered under number MMS/ALU/AE161145—by the Commission Nationale de l’Informatique et des Libertés (CNIL), the French data protection authority. The research was conducted on anonymized data between May 2016 and October 2018, without the possibility of indirect identification. In accordance with the relevant guidelines and regulations, all participants included in the study were informed of and agreed to the possibility of their anonymized data being used for research purposes.

The patient cohort was described using the usual position and dispersion parameters: means and standard deviations or medians and ranges, depending on the nature of the distribution. Categorical data are presented as numbers and percentages.

The primary endpoint was the number of abdominal ultrasounds performed on a patient during each year of follow-up for which the data were available. Surveillance was deemed optimal if the number of ultrasounds was never <2, suboptimal if <2 for one year alone, and failed otherwise (Table 1). Patients for whom follow-up was not available during the year, whatever the reason, were considered to have NA over that year.

**Table 1.** Categorization of surveillance quality according to frequency of ultrasounds over 3-year follow-up period.

| Number of ultrasounds |  |  | Quality of surveillance |
| --- | --- | --- | --- |
| Period 1<br>(months 5 to 16) | Period 2<br>(months 17 to 28) | Period 3<br>(months 29 to 40) |  |
| ≥2 | ≥2 | ≥2 | Optimal |
| ≥2 | ≥2 | NA | Optimal |
| ≥2 | NA | NA | Optimal |
| ≥2 | ≥2 | 1 | Suboptimal |
| ≥2 | 1 | NA | Suboptimal |
| 1 | NA | NA | Suboptimal |
| Any other pattern |  |  | Failed |

NA: Follow-up data not available.

In accordance with our secondary objective of evaluating associations between surveillance level and the sociodemographic or medical variables listed above (see “Data collected”), our primary analysis compared patients with either optimal or failed surveillance, ignoring cases of suboptimal surveillance.

We constructed log-binomial models to measure associations, obtained adjusted measures with a Poisson model adapted to longitudinal data analysis (GEE), and a robust variance estimator to facilitate convergence as well as improve precision, and reported uncertainty in estimates with 95% confidence intervals. Collinearity was checked before introducing the covariates into the multivariate model. All analyses were performed using the SAS software (version 9.4).

## Results

Between January 1, 2009, and December 31, 2013, 75,868 incident cases of cirrhosis were identified (Fig 2). The results are described in Table 2. The mean age of the patients at inclusion was 56 years (SD = 11), and two-thirds of them were male. The initial diagnosis took place in public university hospitals for roughly one-third of the patients and in other public hospitals for half of them. A quarter of the patients benefited from CMU health coverage and were reserved for the most economically disadvantaged in France. Nearly one-tenth of the patients had a Charlson index of 3 or higher.

**Figure 2.**
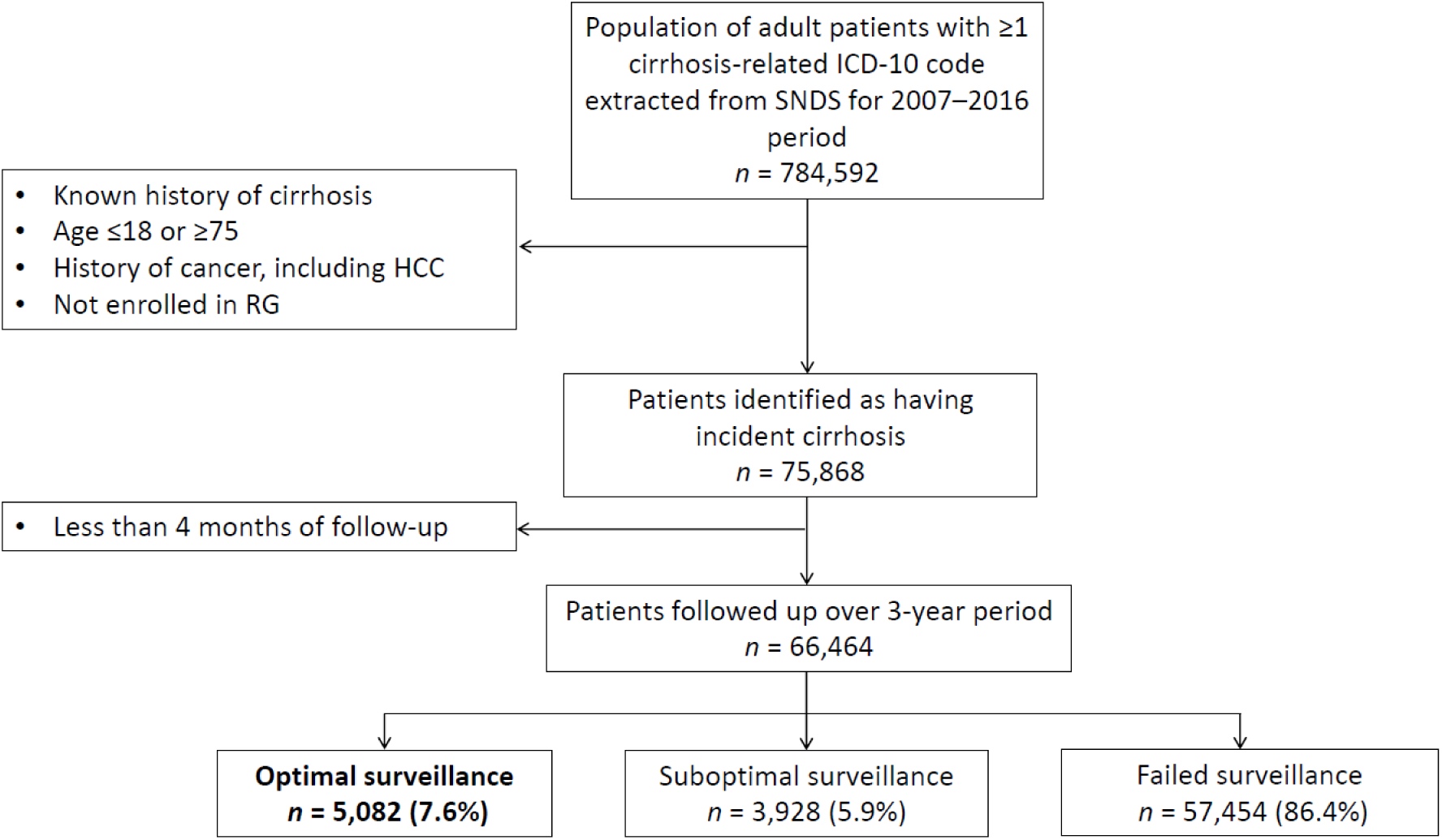
Study flowchart. HCC, hepatocellular carcinoma; RG, Régime Général (French general public health insurance program); SNDS, Système National des Données de Santé (French National Health Data System).

**Table 2.** Characteristics of inception cohort. CMU: Couverture Maladie Universelle (universal health coverage for low-income individuals in France).

|  |  |
| --- | --- |
| **Size of cohort** | 75,868 |
| **Age at inclusion in years, mean (SD)** | 55.6 (11) |
| **Sex, *n* (%)** |  |
| Male | 50,769 (66.9) |
| Women | 25,099 (33.1) |
| **Initial place of care, *n* (%)** |  |
| Public university hospital | 21,457 (28.3) |
| Other public hospital | 37,248 (49.1) |
| Private hospital | 17,163 (22.6) |
| **CMU beneficiary** |  |
| Yes | 18,457 (24.3) |
| No | 57,411 (75.7) |
| **Charlson index** |  |
| 0–1 | 43,999 (58.0) |
| 1–2 | 21,849 (28.8) |
| 3–4 | 7,522 (9.9) |
| ≥5 | 2,498 (3.3) |
| **Etiology** |  |
| Alcoholic | 51,019 (67.2) |
| Viral | 6,736 (8.9) |
| Unknown | 20,758 (27.4) |

Each year, approximately 10% of the patients were censored due to death or a diagnosis of HCC or other cancers (Table 3). Of the 75,868 patients initially identified, 9,404 were censored because they had an ultrasound during the first 4 months, which is more suggestive of diagnostic procedures than surveillance. Of the remaining 66,464 patients, surveillance was optimal in only 7.6% (*n* = 5,082), suboptimal in 5.9% (*n* = 3,928), and failed in 86.4% (*n* = 57,454).

**Table 3.** Characteristics of follow-up.

| Period | Diagnosis<br>(months 1 to 4) | Year 1<br>(months 5 to 16) | Year 2<br>(months 17 to 28) | Year 3<br>(months 29 to 40) |
| --- | --- | --- | --- | --- |
| Number of patients<br>at start of period | 75,868 | 66,464 | 58,647 | 53,192 |
| Patients not censored<br>during follow-up<br>period, *n* (%) | 66,464 (87.6) | 58,647 (88.2) | 53,192 (90.7) | 48,444 (91.1) |
| Number of ultrasounds |  |  |  |  |
| 0 |  | 29,896 (51.0) | 29,372 (55.2) | 28,211(58.2) |
| 1 |  | 15,310 (26.1) | 13,144 (24.7) | 11,481(23.7) |
| ≥2 |  | 13,441 (22.9) | 10,676 (20.1) | 8,752(18.1) |
| Reason for<br>censorship, *n* (%) |  |  |  |  |
| HCC diagnosis | 1,492 (2.0) | 1,232 (1.9) | 882 (1.5) | 763 (1.4) |
| Other cancer<br>diagnosis | 2,270 (3.0) | 1,767 (2.7) | 1,337 (2.3) | 1,198 (2.3) |
| Death | 5,642 (7.4) | 4,818 (7.2) | 3,236 (5.5) | 2,787 (5.2) |

HCC: hepatocellular carcinoma.

Multivariable analysis showed that all variables of interest, other than the type of medical facility, were significantly associated with cirrhosis surveillance. Older age, male sex, high Charlson index, high number of annual visits to GI specialists, and viral etiology were associated with better surveillance aligned with current recommendations, while low socioeconomic status was associated with worse surveillance (Table 4). Sensitivity analysis did not reveal any substantial variation in the results using the alternative comparison pooling optimal and suboptimal surveillance versus failed surveillance.

**Table 4.** Variables associated with optimal HCC surveillance.

| Variable | Univariate analysis,<br>RR (95% CI) | Multivariable analysis,<br>RR (95% CI) |
| --- | --- | --- |
| Age (per 10 years increase) | 1.35 (1.31–1.38) | 1.23 (1.20–1.26) |
| Sex |  |  |
| Female | ref | ref |
| Male | 1.22 (1.16–1.30) | 1.16 (1.10–1.23) |
| Initial place of care |  |  |
| Public university hospital | ref | ref |
| Other public hospital | 0.86 (0.81–0.92) | 0.96 (0.90–1.02) |
| Private hospital | 1.03 (0.96–1.11) | 0.95 (0.88–1.02) |
| Charlson index |  |  |
| 1 | ref | ref |
| 2 | 1.51 (1.43–1.60) | 1.20 (1.13–1.27) |
| 3 | 1.29 (1.18–1.41) | 1.10 (1.00–1.20) |
| 4 | 1.86 (1.65–2.10) | 1.23 (1.09–1.39) |
| Number of hepatologist/gastroenterologist visits per year |  |  |
| 0 | ref | ref |
| 1 | 1.25 (1.02–1.53) | 1.21 (0.99–1.48) |
| >1 | 15.8 (13.7–18.2) | 14.2 (12.2–16.3) |
| Etiology |  |  |
| Alcoholic | ref | ref |
| Viral | 1.88 (1.73–2.05) | 1.30 (1.19–1.42) |
| Low income (CMU beneficiary) |  |  |
| No | ref | ref |
| Yes | 0.67 (0.62–0.72) | 0.92 (0.86–0.98) |

CI: confidence interval; CMU: Couverture Maladie Universelle (universal health coverage for low- income individuals in France); ref: reference value; RR: rate ratio.

## Discussion

In our study, only 13.5% of the 66,464 patients with incident cirrhosis followed-up for 3 years had optimal or suboptimal surveillance, as defined by current European and French guidelines, which recommend semiannual ultrasound for early HCC detection.

Older age, male sex, a high Charlson index, frequent examinations by hepatologists, and viral etiology of the patient’s cirrhosis were identified as factors associated with better surveillance, whereas low socioeconomic status was associated with poorer surveillance.

The low number of patients benefiting from optimal surveillance (7.6%) is consistent with the literature. For example, Yeo et al. [14] reported that in 82,427 American patients followed up for cirrhosis over a mean period of 29 months, between 2007 and 2016, at least one HCC surveillance procedure (ultrasound, CT, or MRI) was performed every 6–12 months for 8.78% of the cases, >12–24 months for 25.32%, and >24 months for 20.47%. Another study, conducted among 597 American patients with hepatitis C virus cirrhosis enrolled in a program at an integrated health system between 2013 and 2020, only 5.0% of patients receiving usual care showed adequate surveillance, defined as at least five surveillance studies within 36 months post-enrollment [21]. In a recent US database nested study conducted on claims ranging from January 2013 to June 2019 among 15,543 patients with cirrhosis, 45.8% and 58.7% had received any abdominal imaging at 6 and 12 months, respectively. Patients were up-to-date with recommended surveillance for only 31% of the median 1.3-year follow-up [22]. In a recent meta-analysis, Wolf et al. reported a global surveillance rate of 24%, varying widely by study location [23]. The surveillance rate in the US (17.8%) was far lower than that in Asia (34.6%).

The factors associated with surveillance have also been linked to surveillance in other countries. Across studies, the number of clinic visits and receipt of care from hepatologists were most consistently and strongly linked to adequate surveillance [23]. Surveillance rates in gastroenterology and hepatology clinics is variable, as high as 75%, but also <10% in large population-based cohorts [23]. In our study, >1 yearly examination by a GI specialist was the strongest driver of optimal surveillance. A positive and independent effect of hepatologist follow-up on the 5-year survival of patients with liver disease has been shown, although patients receiving GI care are highly selected (for more severe cirrhosis and viral etiologies) and may be more inclined to perform a complete follow-up at baseline [24]. Surveillance rates are lower for alcohol etiology than for viral etiology [23]. This observation is of particular importance in France, where alcohol consumption is linked to roughly two-thirds of all HCC cases. Our findings offer new insights for better understanding the poorer prognosis of alcohol-related HCC in France [10,25]. Although we noted an association between older age and optimal surveillance, data from the literature are conflicting [23]. In contrast to other reports [26], we found an association between the Charlson comorbidity index and optimal surveillance. However, one might hypothesize that the impact of comorbidities depends on the severity of the chronic condition in question: providers may be less willing to order surveillance for patients thought to have a lower chance of long-term survival.

The most remarkable finding of our study is the negative impact of low socioeconomic status on the surveillance of patients with cirrhosis, despite the implementation of a system of universal health coverage (CMU) and complete coverage of care expenses for cirrhosis and certain other chronic diseases in France (including transportation). However, patient barriers are not related only to the costs of transport or procedures. Others have been identified, such as travel to the care facility organization, difficulty in scheduling appointments, and identifying where to receive surveillance [27], which brings up the concept of health literacy. Health literacy can be defined as “the personal characteristics and social resources needed for individuals and communities to access, understand, appraise and use information and services to make decisions about health”[28]. Health literacy has been shown to be strongly associated with education, poverty, employment, having a first language different from the national language, and the level of socioeconomic deprivation in the area of residence [29]. Importantly, compared to the rest of the population, CMU beneficiaries are more likely to be blue-collar workers, unemployed, and have lower educational levels. Unfortunately, we were unable to evaluate the impact of language and local socioeconomic deprivation in our study.

However, patients cannot be held accountable for all surveillance failure. Singal et al. showed that physicians frequently failed to order surveillance procedures, especially in patients with alcohol-related cirrhosis [30]. This may in part reflect the persistent controversy concerning the risk-benefit ratio for surveillance in HCC [31,32], despite cumulative evidence in its favor [3,20,33–35], or failure to identify individuals at risk [36]. In addition, the uneven geographical distribution and shrinking numbers of hepatologists and gastroenterologists in France may also play a role, sometimes making physical access to specialized care challenging [37].

Taken together, our results, along with the existing literature, suggest that efforts should be made to improve surveillance programs by addressing these barriers. Several measures to promote HCC surveillance have already been tested with encouraging results [21,23,38–43]. They include patient/provider education as well as both “inreach” (e.g., reminder and recall systems) and outreach (e.g., mailing campaigns to care providers and patients) strategies, each associated with a significant increase in surveillance procedures, albeit heterogeneous in magnitude (+9.4% to +63.6%). Shahsavari et al. and Campbell et al. advocated multilevel interventions with inreach and outreach components that account for individuals’ unique situations in addition to social policies and disparities at the local and national levels [44,45].

A strength of our study was the use of the national SNDS database, which provided a nearly complete picture of medical service consumption by French patients identified as having cirrhosis during the study period.

Our study had some limitations. First, SNDS does not permit the identification of patients with cirrhosis who were not hospitalized during the recruitment period of our study, resulting in an overrepresentation of severe cases in our sample. Second, the SNDS does not provide data on factors such as patient- or physician-reported barriers, or an in-depth clinical profile of each patient that could impact surveillance rates. The results of this study have several implications. First, we may have failed to accurately identify all the cases of incident cirrhosis. Second, the clinical context in which cirrhosis was diagnosed may have prompted the decision not to implement follow-up HCC surveillance.

Another limitation stems from the stringent definition of optimal follow-up, which we chose to be as close as possible to institutional guidelines. First, we did not account for a delay over the six month that defined a period, although an ultrasound occurring shortly afterward may be clinically relevant to declare the period as with surveillance. This may have directly impacted our estimates in a pessimistic manner. Second, we did not consider magnetic resonance imaging or scanners during the periods that could have triggered the decision not to perform ultrasound. It is plausible that a fraction of patients having received recently CT or MRI were falsely identified as not non-compliant with guidelines, although this fraction appeared limited in other similar studies (5%, see Singal et al. [42]). Third, we did not distinguish patients receiving no surveillance at all from patients seldom receiving an ultrasound, which have been classified within the same “failed” surveillance group, although the clinical interpretation of each situation may be different.

Follow-up ended in 2016, well before the COVID-19 pandemic, which is suspected to have had a devastating impact on HCC surveillance [46], albeit transitory [47]. Future interventions aimed at evaluating and promoting surveillance should consider the short- and long-term consequences of the pandemic [48].

In conclusion, our data demonstrated that HCC surveillance in France between 2009 and 2016 was seldom optimal for patients, despite health coverage in France. The greatest predictor of optimal surveillance was frequent examinations by GI specialists. Considering the expected decrease in the number of GI specialists over the next 5–10 years, there is an urgent need to design alternative care pathways to promote and support optimal surveillance in patients with cirrhosis. Equally important, in light of the observed association between low socioeconomic status and failed surveillance, policies aimed at reducing this healthcare disparity need to be implemented, especially knowing that the pandemic has only exacerbated such disparities worldwide.

## Data availability

Data are available from the National Bureau of the French Public Health Insurance Administration (Caisse Nationale de l’Assurance Maladie, or CNAM) for researchers who meet the criteria for access to confidential data set by France’s data protection authority (Commission Nationale de l’Informatique et des Libertés, or CNIL). Authorization was sought from the CNIL (www.cnil.fr) and the French Health Data Hub (www.health-data-hub.fr). This study was registered at ClinicalTrial.gov (NCT02983968).

## Data Availability

Data are available from the National Bureau of the French Public Health Insurance Administration (Caisse Nationale Assurance Maladie, or CNAM) for researchers who meet the criteria for access to confidential data set by french data protection authority (Commission Nationale Informatique et Libertes, or CNIL). Authorization was sought from the CNIL (www.cnil.fr) and the French Health Data Hub (www.health-data-hub.fr).

## Acknowledgements

We thank Jason Miller for language editing and comments that greatly improved the manuscript and Eric Nguyen-Khac, Emmanuel Oger, Jean-Claude Barbare, and Melanie Duval for their useful guidance and proposals.

## Author contributions

Conceptualization: P. C., F. B., N. G.-C., C.C., E. O., O. G. Data curation: F. B.

Formal Analysis: F. B.

Funding acquisition: P. C., O. G. Investigation: P. C., O. G. Methodology: P. C., F. B., E. O., O. G.

Project administration: P. C., E. O., O. G. Resources: P. C., O. G.

Software: F. B.

Supervision: P. C., E. O. O. G.

Validation: P. C., F. B., E. O. O. G. Visualization: P. C.

Writing – original draft: P. C., N. G.-C., F. B., E. O., C. C., O. G.

Writing – review & editing: P. C., N. G.-C., F. B., E. O., C. C., O. G.

---

**S1 Table.**
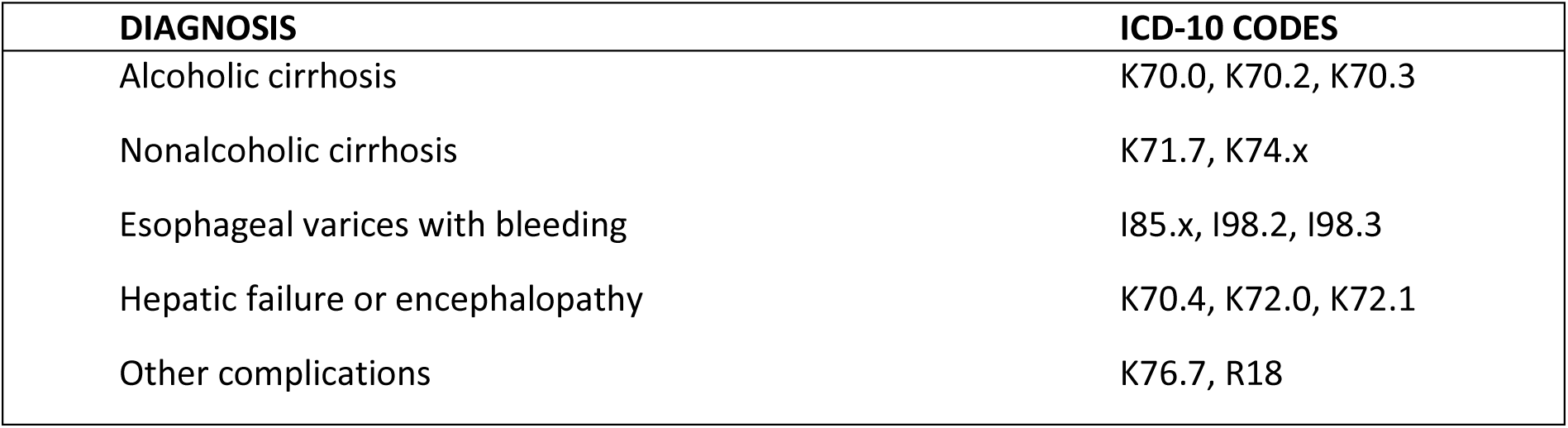
ICD-10 codes used to identify cirrhosis in the SNDS database.

| DIAGNOSIS | ICD-10 CODES |
| --- | --- |
| Alcoholic cirrhosis | K70.0, K70.2, K70.3 |
| Nonalcoholic cirrhosis | K71.7, K74.x |
| Esophageal varices with bleeding | I85.x, I98.2, I98.3 |
| Hepatic failure or encephalopathy | K70.4, K72.0, K72.1 |
| Other complications | K76.7, R18 |

**S2 Table.** French CCAM medical procedure codes for identification of liver Doppler ultrasounds.

| CCAM CODE | DESCRIPTION |
| --- | --- |
| ZCQM001 | Transcutaneous ultrasound of the abdomen, with Doppler ultrasound of the digestive vessels |
| ZCQM002 | Transcutaneous ultrasound of the abdomen, with transcutaneous ultrasound of the pelvis and Doppler ultrasound of the digestive vessels |
| ZCQM004 | Transcutaneous ultrasound of the upper abdomen with Doppler ultrasound of the digestive vessels |
| ZCQM005 | Transcutaneous ultrasound of the abdomen, with transcutaneous ultrasound of the pelvis |
| ZCQM006 | Transcutaneous ultrasound of the upper abdomen |
| ZCQM008 | Transcutaneous ultrasound of the abdomen |
| ZCQM010 | Transcutaneous ultrasound of the upper abdomen and pelvis |
| ZCQM011 | Transcutaneous ultrasound of the upper abdomen and pelvis with Doppler ultrasound of the digestive vessels |
| HLQM001 | Transcutaneous ultrasound of the liver and bile ducts |

